# Discovering additional genetic loci associated with six psychiatric disorders/traits *via* FDR regression model leveraging external genetic and biological data

**DOI:** 10.1101/2024.01.29.24301912

**Authors:** Shi-tao Rao, Jing-hong Qiu, Yi-qiang Zhi, Yu-ping Lin, Ruo-yu Zhang, Xiao-tong Chen, Dan Xu, Hon-Cheong So

**Affiliations:** Department of Bioinformatics, Fujian Key Laboratory of Medical Bioinformatics, Institute of Precision Medicine, School of Medical Technology and Engineering, Fujian Medical University, Fuzhou, Fujian, China; School of Biomedical Sciences, The Chinese University of Hong Kong, N.T., Hong Kong; School of Basic Medical Sciences, Fujian Medical University, Fuzhou, Fujian, China; Fujian Key Laboratory of Molecular Neurology, Institute of Neuroscience, Fujian Medical University, Fuzhou, Fujian, China; KIZ-CUHK Joint Laboratory of Bioresources and Molecular Research of Common Diseases, Kunming Institute of Zoology and The Chinese University of Hong Kong, Kunming, China; CUHK Shenzhen Research Institute, Shenzhen, China; Department of Psychiatry, The Chinese University of Hong Kong, Shatin, N.T., Hong Kong; Margaret K.L. Cheung Research Centre for Management of Parkinsonism, The Chinese University of Hong Kong, Shatin, N.T., Hong Kong; Brain and Mind Institute, The Chinese University of Hong Kong, Shatin, N.T., Hong Kong; Hong Kong Branch of the Chinese Academy of Sciences (CAS) Center for Excellence in Animal Evolution and Genetics, The Chinese University of Hong Kong, Shatin, N.T., Hong Kong

## Abstract

**Background:** Common psychiatric disorders have substantial heritability influenced by multiple genes. While a number of susceptibility variants have been identified, many associated variants remain undiscovered. This study aimed to identify additional genetic loci associated with common psychiatric disorders/traits by leveraging correlated traits and biological annotations.

**Methods:** We proposed application of the false discovery rate (FDR) regression model to uncover additional genetic loci for six psychiatric disorders/traits. To enhance the likelihood of discovering additional significant genetic loci and genes, we utilized a set of 42 correlated traits and 21 biological annotations as covariates. Internal validation analysis and drug cluster enrichment analysis were conducted to validate the biological significance of the additional genetic loci/genes uncovered. We also experimentally validated two additional genes revealed for autism spectrum disorder (ASD).

**Results:** The FDR regression (FDRreg) analysis strategy revealed hundreds of additional significant genes (FDR<0.01) in gene-level analyses, surpassing the number of significant genes found in the original studies. Specifically, in 11/16 trait analyses, FDRreg identified more significant genes based on gene-based analysis with MAGMA, and in 12/16 analyses, FDRreg identified more significant genes based on imputed expression in the brain. In SNP-level results, the majority of analyses (13/16) identified an equal or higher number of genomic risk loci (FDR<0.01). We found that FDRreg is able to reveal genes that are later known to be significant in subsequent larger-scale GWAS. Drug cluster enrichment analysis demonstrated a stronger enrichment in psychiatry-related drug clusters. In utero electroporation (IUE) experiments provided evidence to support two additional genes identified for ASD in critical embryonic brain development processes.

**Conclusions:** By integrating genetically correlated traits and biological annotations, the FDRreg strategy enables the identification of a greater number of additional significant genes and risk loci. Moreover, the new associated genes exhibited meaningful biological and clinical implications. This study presents a valuable approach for uncovering the genetic basis of psychiatric disorders and gaining insights into their underlying biology.

## Introduction

Although most psychiatric disorders are reported to have a high heritability, the genetic basis remains unclear [1]. Currently, the genetic basis of psychiatric disorders is considered highly polygenic, with each genomic variant explaining only a small portion of the heritability. The genome-wide association study (GWAS) strategy is widely used to identify associated variants for complex human diseases, as it allows for the simultaneous analysis of thousands of variants [2, 3]. However, due to the small effect size of each variant, large sample sizes are required to confirm the true association signals. Fortunately, with the increasing availability of GWAS summary statistics from various international consortia studying the etiology of psychiatry disorders, it is now possible to significantly increase the sample size and apply advanced methods to analyze this vast amount of data. This approach greatly enhances the detection power and enables the identification of true association signals for diseases of interest [4]. In comparison to using individual-level data, GWAS summary statistics can be easily obtained from publicly released sources [3].

At the outset of employing the GWAS strategy, studies were typically conducted for individual traits, which suffered from a relatively low statistical power [5]. To address this limitation, multivariate analysis strategies have been developed to simultaneously analyze multiple traits. However, most multivariate methods still require individual-level data for both genotypes and phenotypes [5]. On the other hand, alternative methods were proposed to integrate results from univariate analyses across various phenotypes. These methods included Fisher’s combined p-value method, standard fixed-effects and random-effects meta-analysis methods, as well as several statistical methods based on a linear combination of univariate test statistics [6–10]. While these methods have identified more risk loci or associated genes, they have also introduced potential issues. For instance, aggregating p-values of different but correlated phenotypes within the same cohort could result in an inflated type I error [4, 11]. Furthermore, these methods generally do not allow for heterogeneity across cohorts or situations where genetic variants have opposite effects on different phenotypes [8]. More recently, the cross-phenotype association (CPASSOC) and multi-trait analysis of GWAS (MTAG) methods have been developed to address some of these issues [4, 12]. However, certain challenges still remain. For example, while MTAG is computationally efficient in generating trait-specific effect estimates for each SNP when applied to multiple traits, the computation time for maximum false discovery rate (FDR) increases exponentially when including numerous traits. This hampers the necessary validation step for the associated signals [12]. Additionally, MTAG assumes that all SNPs share the same variance-covariance matrix of effect sizes across traits, which could be problematic for SNPs that are truly null for one trait but non-null for another, leading to an increased rate of false positives [12]. On the other side, the CPASSOC method is primarily considered as a meta-analysis approach that utilized summary-level data from single SNP-trait associations in GWAS to detect variants associated with at least one trait [4].

Another potential issue is the difficulty in interpreting the biological function of associated variants[3]. To date, most of the identified association variants are located in intergenic or intronic regions, which may not directly impact the expression or structure of functional proteins [3]. This phenomenon hinders the design of suitable experiments to validate the underlying mechanisms of the associations and elucidate the pathophysiology of diseases [13]. Additionally, conducting direct functional validation for the numerous potentially causal variants or genes requires expensive and labor-intensive experiments [13].

Here we presented an analytic framework based on the FDR regression (FDRreg) approach to identify additional association signals by integrating association evidence from GWAS summary statistics of related traits/disorders, and a wealth of biological information, which may be derived from real experimental data (**Fig. 1**). For instance, the regression model could incorporate evidence from drug targets or animal models. This statistical method not only effectively controlled the false discovery rate, but also extracted association evidence from multiple correlated traits related to the target phenotype. This significantly increased the likelihood of discovering novel risk loci or associated genes/pathways.

In brief, the proposed analytic framework and study based on FDRreg differs from previous approaches in several ways and has the following strengths: (1) The proposed framework is designed to output test statistics for the *primary* trait/study of interest, unlike meta-analysis in which the objective is to combine evidence across all relevant traits/studies. For example, if one is interested in the genetic associations of schizophrenia (SCZ) only but wish to leverage GWAS results from bipolar disorders (BD) and depression, our approach can output the significance of SNPs/genes pertaining to SCZ only, with consideration of genetic evidence from related traits. Meta-analysis on the other hand produces test statistics by *aggregating* associations from all three traits, and the interpretation will be different; (2) The method does not require restrictive assumptions as in MTAG (e.g. assuming all SNPs share the same variance-covariance matrix of effect sizes across traits); (3) it can potentially handle GWAS results of related traits that are heterogeneous, as FDRreg will estimate the prior probabilities of associations adaptively, based on how strong the covariates are linked to the significance of the primary trait. Even if some irrelevant traits are included, they tend to have only minor contributions to the final results; (4) The computational speed is relatively fast and can scale to large-scale GWAS (or other omics) datasets; (5) as discussed above, we can incorporate ***both*** GWAS results of related traits and biological information simultaneously for statistical inference, which cannot be achieved by meta-analysis approaches or MTAG; (6) At the same time, we could extract which biological features contribute to significant associations (and which may be irrelevant), providing important scientific insights; (7) In addition to the above, we performed further functional studies (in utero electroporation, IUE) to verify the role of several selected genes revealed by our analytic framework. Experimental validation provides further support to the validity and potential of our proposed method in yielding new discoveries. In addition, the IUE study helped to elucidate the functional role of the selected genes in neurodevelopment, and the findings are of interest in its own right.

In the current study, we applied the strategy to investigate six different psychiatric disorders/traits. This approach successfully identified hundreds of additional significant genes (FDR<0.01, **Table 1**) in gene-level analyses, surpassing the original results. The integration of association evidence from multiple correlated traits and biological annotations played a crucial role in this discovery. Furthermore, we observed that for the majority of analyses (13/16, **Table 1**), we identified either an equal number or more genomic risk loci compared to the original findings (without FDRreg). To validate the biological relevance of the identified genes, we conducted internal validation, by testing whether the new genes uncovered by our approach will later turn out to be significant in subsequent larger-scale GWAS (**Table 2**). Additionally, the drug cluster enrichment analysis revealed significant enrichment of associated signals from FDRreg analysis in psychiatry-related drug clusters, such as antidepressants, antipsychotics and dopaminergic agents (**Table 3**). Beyond psychiatric disorders, this proposed strategy holds great potential for discovering additional risk loci or associated genes in other complex diseases.

**Table 1.**
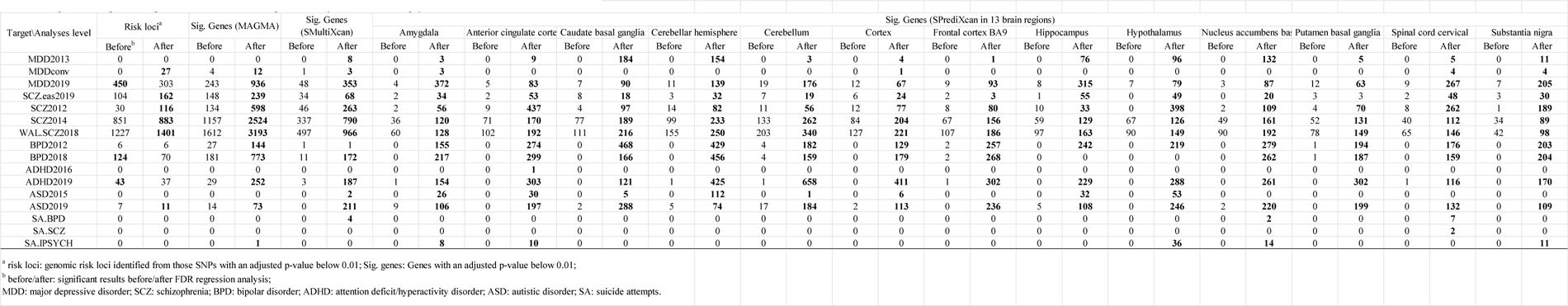
Comparison of significant signals with or without FDR regression analysis for six kinds of psychiatric disorders/outcome.

**Table 2.** Internal validation analysis based on significant genes from small-size and big-size GWAS data.

|  |  |  |  |  |  |  |  |  |  |  |  |  |
| --- | --- | --- | --- | --- | --- | --- | --- | --- | --- | --- | --- | --- |
| overlap (b): number of significant genes that overlapped between small-size study (results after FDRreg analysis) and big-size study (original results).<br>overlap (a): number of significant genes that overlapped between small-size study (original results) and big-size study (original results).<br>fold change: overlap(b)/overlap(a); x: no. of significant genes from big-size study (original results).<br>p.value: caculated by proportion test or by fisher's exact test in case of small number in cells.<br>SCZ: schizophrenia; BPD: bipolar disorder; MDD: major depressive disorder. |  |  |  |  |  |  |  |  |  |  |  |  |
| Program/comparison | SCZ2012 vs. WAL.SCZ2018 |  |  |  | BPD2012 vs. BPD2018 |  |  |  | MDD2013 vs. MDD2019 |  |  |  |
|  | overlap(a) | overlap(b) | fold change | x | overlap(a) | overlap(b) | fold change | x | overlap(a) | overlap(b) | fold change | x |
| MAGMA | 305 | 852 | **2.79** | 2654 | 44 | 131 | **2.98** | 482 | 0 | 0 | NA | 578 |
| SMultiXcan | 127 | 285 | **2.24** | 1004 | 2 | 1 | 0.50 | 120 | 7 | 14 | **2.00** | 200 |
| SPrediXcan (13 brain regions) |  |  |  |  |  |  |  |  |  |  |  |  |
| Amygdala | 9 | 44 | **4.89** | 110 | 0 | 2 | **Inf** | 2 | 0 | 0 | NA | 7 |
| Anterior_cingulate_cortex_BA24 | 13 | 89 | **6.85** | 171 | 0 | 7 | **Inf** | 12 | 0 | 1 | **Inf** | 20 |
| Caudate_basal_ganglia | 16 | 74 | **4.63** | 209 | 1 | 8 | **8.00** | 15 | 0 | 7 | **Inf** | 33 |
| Cerebellar_Hemisphere | 22 | 91 | **4.14** | 266 | 1 | 10 | **10.00** | 23 | 0 | 4 | **Inf** | 27 |
| Cerebellum | 31 | 82 | **2.65** | 359 | 4 | 18 | **4.50** | 35 | 0 | 0 | NA | 37 |
| Cortex | 17 | 72 | **4.24** | 235 | 0 | 9 | **Inf** | 22 | 0 | 0 | NA | 26 |
| Frontal_Cortex_BA9 | 12 | 59 | **4.92** | 177 | 1 | 9 | **9.00** | 16 | 0 | 2 | **Inf** | 31 |
| Hippocampus | 10 | 39 | **3.90** | 159 | 0 | 3 | **Inf** | 8 | 0 | 6 | **Inf** | 19 |
| Hypothalamus | 9 | 67 | **7.44** | 155 | 0 | 3 | **Inf** | 5 | 0 | 4 | **Inf** | 16 |
| Nucleus_accumbens_basal_ganglia | 6 | 65 | **10.83** | 165 | 1 | 6 | **6.00** | 11 | 0 | 2 | **Inf** | 19 |
| Putamen_basal_ganglia | 12 | 53 | **4.42** | 166 | 1 | 4 | **4.00** | 9 | 0 | 0 | NA | 21 |
| Spinal_cord_cervical_c-1 | 10 | 53 | **5.30** | 104 | 0 | 4 | **Inf** | 4 | 0 | 1 | **Inf** | 14 |
| Substantia_nigra | 5 | 46 | **9.20** | 83 | 0 | 2 | **Inf** | 3 | 0 | 0 | NA | 11 |

**Table 3.** Comparison of enriched top 7 psychiatry-related drug clusters before or after FDR regression analysis for SCZ, MDD and BPD.

| Drug cluster\Disorder | Code | SCZ2012 |  |  |  | SCZ2014 |  |  |  | WAL.SCZ2018 |  |  |  | BPD2012 |  |  |  |
| --- | --- | --- | --- | --- | --- | --- | --- | --- | --- | --- | --- | --- | --- | --- | --- | --- | --- |
|  |  | P(before)<sup>a</sup> | FDR(before) | P(after)<sup>b</sup> | FDR(after) | P(before)<sup>a</sup> | FDR(before) | P(after)<sup>b</sup> | FDR(after) | P(before)<sup>a</sup> | FDR(before) | P(after)<sup>b</sup> | FDR(after) | P(before)<sup>a</sup> | FDR(before) | P(after)<sup>b</sup> | FDR(after) |
| Anti-epileptics | N03A | 2.90E-04 | **2.20E-02** | 1.91E-06 | **1.45E-04** | 2.14E-04 | **2.51E-03** | 2.74E-04 | **7.10E-03** | 1.83E-04 | **1.39E-02** | 2.79E-04 | **8.48E-03** | 1.26E-01 | 8.79E-01 | 1.41E-05 | **1.08E-03** |
| Antidepressants | N06A | 6.24E-03 | 2.01E-01 | 4.09E-06 | **2.07E-04** | 1.75E-01 | 3.85E-01 | 1.15E-03 | **2.18E-02** | 2.86E-02 | 2.09E-01 | 1.13E-03 | **1.99E-02** | 2.98E-01 | 1.00E+00 | 1.12E-03 | **4.27E-02** |
| Antipsychotics | N05A | 3.14E-05 | **4.77E-03** | 4.71E-10 | **7.15E-08** | 2.04E-07 | **3.11E-05** | 1.13E-10 | **1.72E-08** | 1.70E-09 | **2.58E-07** | 1.31E-12 | **1.99E-10** | 5.08E-02 | 5.55E-01 | 9.75E-11 | **1.49E-08** |
| Dopaminergic agents | N04B | 6.22E-02 | 3.32E-01 | 9.66E-05 | **2.94E-03** | 1.67E-02 | 6.87E-02 | 9.01E-04 | **1.96E-02** | 2.25E-03 | 8.55E-02 | 1.18E-04 | **7.93E-03** | 3.89E-01 | 1.00E+00 | 6.90E-05 | **3.52E-03** |
| Psychostimulants | N06B | 2.19E-01 | 4.66E-01 | 1.01E-01 | 4.18E-01 | 2.81E-01 | 5.57E-01 | 1.23E-01 | 3.00E-01 | 1.38E-01 | 4.06E-01 | 7.84E-02 | 3.85E-01 | 4.86E-01 | 1.00E+00 | 1.44E-01 | 6.13E-01 |
| Anxiolytic | N05B | 6.14E-01 | 8.23E-01 | 6.50E-03 | 8.98E-02 | 6.08E-01 | 9.15E-01 | 5.41E-02 | 2.39E-01 | 3.60E-01 | 6.83E-01 | 8.75E-02 | 3.91E-01 | 4.80E-02 | 5.55E-01 | 8.04E-02 | 5.06E-01 |
| Hypnotics | N05C | 4.85E-02 | 3.21E-01 | 2.12E-05 | **8.05E-04** | 7.49E-03 | **3.67E-02** | 3.85E-05 | **1.95E-03** | 3.59E-02 | 2.09E-01 | 1.62E-04 | **7.93E-03** | 2.01E-01 | 1.00E+00 | 5.25E-02 | 5.02E-01 |

<sup>a</sup> P(before/after): nominal p-value before FDR regression analysis and 'equivalent' nominal p-value after FDR regression analysis for all genes from genome-wide gene-based association study.
<sup>b</sup> FDR(before/after): adjusted p-value before or after FDR regression analysis for all genes from genome-wide gene-based association study.
SCZ: schizophrenia, BPD: bipolar disorder, MDD: major depressive disorder.
Adjusted p-values below 0.05 are in bold.

## Materials and Methods

### 2.1 Target psychiatric disorders and relevant phenotypes

In this study, our focus was on five common psychiatric disorders: major depressive disorder (MDD), schizophrenia (SCZ), bipolar disorder (BPD), autism spectrum disorder (ASD), attention deficit/hyperactivity disorder (ADHD) and one psychiatric outcome, suicide attempt (SA). We obtained GWAS summary statistics for these targets from various sources, including the psychiatric genomics consortium (PGC), the integrative psychiatric research (iPSYCH) project, and the Centre for Neuropsychiatric Genetics and Genomics, respectively (**Supplementary Table 1.1, Table S1.1**). Additionally, we searched for potentially relevant phenotypes from freely accessible GWAS summary statistics repositories. These phenotypes were included as variables in the FDR regression model to provide as much genetic information as possible for the corresponding targets. Overall, we identified a total of 42 GWAS datasets that were included as model variables in the study (**Table S1.2**). The latest available GWAS data of these phenotypes were employed to maximize the amount of genetic information utilized in the analysis.

### 2.2 Initial quality controls and variables selection

To ensure the quality of the selected summary statistics, we conducted initial quality control steps. This involved filtering out duplicated SNPs and insertion/deletion (InDel) variants from the datasets. Next, we utilized the DIST program to impute summary statistics of unmeasured SNPs with the 1000 Genomes reference population panel [14]. SNPs with an imputation quality score below 0.5 were excluded from further analysis. For the cleaned datasets, we assessed the genetic correlation for each pair of related trait and target (i.e., our primary disorder of interest) using the Linkage-Disequilibrium SCore regression (LDSC) program [15]. This analysis utilized a dataset containing over 1,217,000 SNPs (HapMap3) as a reference for measuring genetic correlations. A p-value below 0.05 was considered indicative of a significant genetic correlation between a trait and the target phenotype. For traits that showed significant genetic correlation, we performed a harmonization step to ensure the signed statistics of the reference allele remained consistent between each pair of correlated trait and target. This was accomplished using the ‘harmonise_data’ function in the ‘TwoSampleMR’ package in R 3.5.3 [16]. Additionally, we conducted a decorrelation step for pairs of traits with sample overlap, to address any potential bias in FDR estimation due to correlation of test statistics owing to overlap. A decorrelated Z-score matrix was generated based on LeBlanc *et al.* [17], where C is a matrix with ones at the diagonal and the LDSC intercept [20] serving as sample overlap correlation coefficients at the other positions, and matrix Z contains the original Z-scores.

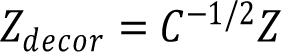

### 2.3 Functional annotations for whole-genome genes

For the annotations of mapped genes, we utilized the Database for Annotation, Visualization, and Integrate Discovery (DAVID, v6.8) to map biological information to each gene [18]. From this database, we selected six types of information: expression-specific tissues, associated diseases, relevant pathways, transcription factor binding sites (TFBS), biological processes, and biological interactions. Additionally, we incorporated biological information from the Open Targets database, which provides associations of each gene with five common psychiatric disorders (SCZ, BPD, MDD, ASD, ADHD). This included information such as drug target, animal model, and literature support of drug targets [19]. Furthermore, we searched the denovo database (denovo-db, version 1.6.1) to identify if a gene had de novo mutation observed in either SCZ or ASD patients [accessed 02-2020]). Overall, we selected a total of 21 correlated biological annotations from these three databases to provide comprehensive biological information for whole-genome genes. For more details about these annotations, please refer to the supplementary notes.

### 2.4 FDR regression analysis for whole-genome SNP-based and gene-based association studies

In the present study, we utilized the false discovery rate (FDR) regression model to adjust the significance level of all SNPs obtained from the GWAS summary data of the primary trait. This model can incorporate information from other GWAS summary data of genetically related traits and relevant biological factors.

We highlight the principles of FDR regression here; please refer to the original paper [20] for details. For test statistics *z*, assume π_1_ is the (prior) probability of being non-null, and *f_1_*(*z*) and *f_0_*(*z*) are distributions of *z* under H_1_ and H_0_ respectively. We may assume a mixture model as follows

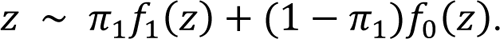

The local false discovery rate (local fdr) is Prob(H_0_|*z*), i.e. 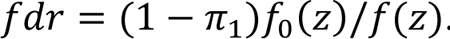. Here we also incorporate external data (***x***, which can be a single or a vector of covariates), such that the model becomes

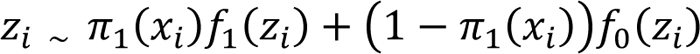

In this case, the local fdr or FDR are modified accordingly with new estimates of π_1_, which is a function of the covariates. The FDRreg program employs a logistic model to model π_1_(*x_i_*),

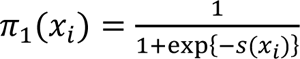

where 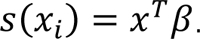.

Note that in FDRreg, the distribution of z under the null and alternative are assumed to follow

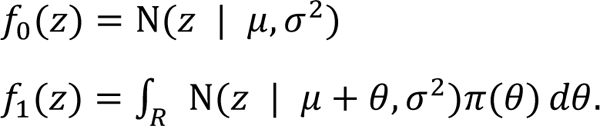

In the SNP-based regression model, the target GWAS *z-*score statistics were considered as the outcome variable. The absolute values of GWAS summary statistic *z*-score from other related traits were treated as model covariates. The null type ‘theoretical’ was chosen, as it was shown to produce more stable estimates than the empirical null [20]. Following the FDR regression analysis, SNPs with an FDR<0.05 were considered statistically significant. These significant SNPs were then utilized for the identification of genomic risk loci using the PLINK program (version 1.9). The following parameters were employed for clumping: a significance threshold of p-value=0.01 for index SNPs, a secondary significance threshold of p-value=0.1 for clumped SNPs, an LD threshold of r^2^=0.01, and a physical distance threshold of distance=1000kb.

For the gene-based association study, we first utilized the ‘MAGMA’ program to conduct whole-genome gene-based association studies on the summary statistics [21]. From these analyses, we would calculate gene-based *z-*score statistics based on the obtained p-values. The covariates consisted of a combination of *z*-scores from the summary statistics of related traits and covariates representing biological relevance of the corresponding genes. Briefly, the significantly correlated traits identified from the genetic correlation analysis (*P*< 0.05) were selected as covariates for the regression model. Additionally, all the matched biological annotations with the corresponding genes were added into the model to modify the significance level of genes.

### 2.5 FDR regression analysis for imputed gene expression

For both target and variable summary statistics, we initially conducted gene expression imputation analyses in 13 different functional brain regions respectively using the ‘S-PrediXcan’ function within the ‘MetaXcan’ program [22]. These brain regions encompassed the amygdala, anterior cingulate cortex (BA24), caudate basal ganglia, cerebellar hemisphere, cerebellum, cortex, frontal cortex (BA9), hippocampus, hypothalamus, nucleus accumbens (basal ganglia), putamen (basal ganglia), spinal cord (cervical c-1), and substantia nigra. The gene-based expression imputation *z-scores* were then directly added into the regression model. Subsequently, FDR regression analyses were performed separately for each brain region. Additionally, the ‘S-MultiXcan’ function within the program allowed us to integrate the gene expression imputation analyses across the 13 brain regions, using GWAS summary statistics. The generated *z-scores* from p-values were directly incorporated into the model, enabling the FDR regression analysis to be conducted for integrated brain regions as well.

### 2.6 Internal validation and functional annotations

To assess the detection capacity and accuracy of outcomes generated from the FDR regression analyses, internal validations were conducted. This process involved determining the number of overlapping genes between significant genes from FDR regression analysis for a small sample set (e.g. BPD2012) and the original significant genes for a larger sample set (e.g. BPD2018) (‘a’ and ‘x’, **Table 1.3**). Similarly, the number of genes between original significant genes for a small sample set (e.g. BPD2012) and that for a larger sample set (e.g. BPD2018) were also calculated (‘b’ and ‘x’, **Table 1.3**). The genes discovered in the larger-scale GWAS are considered as a “standard”, and we hypothesize that FDRreg is able to detect a larger number of genes later turned out to be significantly associated with the disease (i.e., a is larger than b).

In addition to the internal validations, we conducted gene ontology (GO) set and pathway enrichment analysis for the significant genes identified, using the ConsensusPathDB database (CPDB, human) (http://consensuspathdb.org/) [23]. This database contains pre-defined pathway-based sets from various pathway databases, such as KEGG, Reactome, Wikipathways and others. It also includes GO-based sets that are annotated with specific GO terms. Furthermore, we performed drug enrichment analysis for the significant genes to identify enriched drugs from a dataset of 17,840 types of drugs. The significantly enriched drugs were then subject to drug cluster analysis from a dataset of 267 drug clusters. These methods have been employed in our previous studies [24].

### 2.7 Phenotypic study of candidate genes in embryonic brain development

To investigate the biological function of additional association genes identified for ASD using the proposed FDR regression model, we primarily focused on the intersected ASD-associated genes resulting from FDR regression analysis across three brain regions, including anterior cingulate cortex BA24, cortex, and frontal cortex. Furthermore, all intersected genes underwent evaluation to determine if they exerted a discernible biological function, primarily based on accumulated evidence available in the OpenTarget database. This assessment involved examining whether the gene in question is a pseudogene or encodes a biologically active protein. Additionally, we examined their expression level in different stages of central nerve system development and various organs in adults. Subsequently, we conducted in utero electroporation (IUE) experiments to identify the biological functions of genes in rat brain at embryonic day (E) 16.5, with tissue harvested at E20.5.The E16.5 rats were chosen for IUE based on previous protocols [25]. All animal procedures performed in this study were conducted in compliance with the protocols approved by the Institutional Animal Care and Use Committee of Fujian Medical University.

To assess the knockdown capacity of ASD-associate genes, we screened bicistronic constructs that encoded both EGFP and shRNA in 293T cell lines. The cell lines were cultured in DMEM with 10% FBS. Transfection and western blotting were performed following previously established protocols [25]. Subsequently, using the selected highly effective shRNAs, we depleted the expression of candidate genes in neural progenitor cells (NPCs) *via* IUE at E16.5 rat brains. We then observed the changes in expression four days later at E20.5. For cryosections, the harvested tissues were fixed in 4% PFA, cryoprotected in 30% sucrose, and frozen in tissue freezing medium. Immunofluorescence staining was conducted in a manner similar to previously described methods [26]. For more detailed information regarding the antibodies used and DNA constructs, please refer to the supplementary notes.

## Results

Please note that all supplementary tables are available at https://drive.google.com/drive/folders/1xFh13JsIEVoJ1jeKv2Og6HAvQGQzaqqk?usp=sharing

### FDR regression analyses for six psychiatric disorders/outcome

For each psychiatric disorder/outcome, the FDR regression analysis strategy was applied to more than one target GWAS data sets with different sample sizes or populations (**Table S1.1**). For instance, both the GWAS studies of BPD published in 2012 and 2018, with significantly different sample sizes, were considered as targets. In total, 16 different GWAS datasets were included as targets in the present study (**Table S1.1**). By combining various genetic and biological factors as co-variables in FDR regression model, we identified a significantly higher number of significant genes (FDR<0.01) for most of gene-level analyses compared to the original results. Specifically, 11 out of 16 analyses for significant genes from the MAGMA program and 12 out of 16 for significant genes from the S-MultiXcan program showed an increase in the number of significant genes (**Table 1**). The remaining analyses showed an equal or similar number of significant findings before and after the FDR regression analysis. In terms of SNP-level results, only genetic factors were considered as co-variables and added into the FDR regression model. As a result, the majority of the analyses (13/16) identified a higher or equal number of genomic risk loci (FDR<0.01, **Table 1**).

#### Schizophrenia

For SCZ, we selected four GWAS datasets as targets due to their varying sample size (SCZ2012, SCZ2014, WAL.SCZ2018 and SCZ.eas2019; **Table S1.1**). Additionally, one of these datasets (SCZ.eas2019) specifically focused on the East Asian population rather than the European population, making it a unique dataset for SCZ. After conducting genetic correlation analyses of these targets with the library of 42 co-variables (GWAS results of related neuropsychiatric traits), we found that 27 co-variables were significantly associated with SCZ2012, 33 co-variables with SCZ2014, 32 co-variables with WAL.SCZ2018, and 20 co-variables with SCZ.eas2019 (**Table S2.1, S3.1, S4.1 and S5.1**).

With the additional information obtained from the genetically correlated GWAS datasets and biological factors, the results generated after the FDR regression analyses for all four targets were highly promising (**Table 1**). At the genomic loci level, the FDR regression analysis was able to identify a significantly higher number of risk loci for all four targets (**Table 1**). At the gene level, the methods employed were able to uncover more significantly associated genes in most of the analyses for the whole-genome gene-level association study, imputed gene expression studies for the integrated results of 13 brain regions, and for all the individual brain regions (FDR<0.01, **Table 1**). For detailed information on the associated risk loci/genes, please refer to Table S2 through Table S5.

To validate the biological significance of these results, we conducted an internal validation analysis. We observed that the number of overlapped genes between the significant genes identified after the FDR regression analysis for SCZ2012 and the original significant results from WAL.SCZ2018 was substantially higher than that between the original significant results of SCZ2012 and WAL.SCZ2018 (**Table 2, Table S4.4**). Similarly, the comparison between SCZ2012 and SCZ2014 also yields promising results (**Table S3.4**). These findings indicated that the FDR regression methods were able to identify additional genes that are likely truly linked to the disorder.

In addition to the internal validation, we also conducted pathway and GO set enrichment analyses for the genome-wide significant genes identified after the FDR regression analysis of WAL.SCZ2018, as it contained the largest number of significant genes (**Table S4**). The pathway enrichment analysis for these genome-wide associated genes suggested significant enrichment in neuron/synapse related pathways, including neuronal system (FDR=7.24E-13, **Table S4.5**), alcoholism of human (FDR=4.80E-10), transmission across chemical synapses (FDR=5.57E-09), and neurotransmitter receptors and postsynaptic signal transmission (FDR=7.14E-07). Moreover, the GO sets enrichment analysis identified neuron/synapse related significantly enriched sets, such as generation of neurons (FDR=3.59E-28, **Table S4.5**), anterograde trans-synaptic signaling (FDR=2.13E-20), and regulation of nervous system development (FDR=3.05E-18). Furthermore, these genes were significantly enriched in several novel pathways, such as WNT signaling related pathways and Rho GTPase activating proteins. Additionally, individual drug and drug cluster enrichment analyses were performed for these significant genes, and significant enrichment was observed across various drugs (**Table S4.7**). Comparing the identified significant genes after the FDR regression analysis with the original associated signals for all the three SCZ targets (SCZ2012, SCZ2014 and WAL.SCZ2018), the significant genes showed stronger enrichment signals in five psychiatry-related clusters: anti-epileptics, antidepressants, antipsychotics, dopaminergic agents, and hypnotics (**Table 3**; more details in **Table S2.9, S3.10** and **S4.9**).

#### Major depressive disorder

For MDD, three different GWAS datasets were selected because they had different sample sizes or were conducted in different populations (European and Asian, **Table S1.1**). Among the library of 42 co-variables, 26 co-variables were identified to be genetically associated with MDD2013, 10 co-variables with converge MDD (MDDconv), and 34 co-variables with MDD2019 (p-value<0.05, **Table S6.1, S7.1** and **S8.1**).

Regarding MDD2013, after the FDR regression analysis, we identified eight additional significant genes for the integrated results of 13 brain regions (FDR<0.01, **Table 1**). Moreover, there were also more significant genes identified for all the 13 individual brain regions compared to their original results (**Table 1**). For MDDconv, the 10 associated genetic co-variables and 21 biological factors provided substantially additional information, resulting in a substantial increase in the number of genomic risk loci from 0 to 27 (**Table 1**). Furthermore, the FDR regression analysis identified more significant genes in the whole-genome gene-level association study, imputed gene expression studies (integrated results of 13 brain regions), and for four individual brain regions (amygdala, cortex, spinal cord cervical, and substantia nigra). As for MDD2019, the analysis did not identify additional risk loci, but it did discover a significantly higher number of significant genes in all gene-level analyses (FDR<0.01, **Table 1**). For detailed information on the associated risk loci/genes, please refer to Table S6 through S8.

To validate the results from FDR regression analysis, an internal validation analysis was conducted to examine if the significant genes identified by the analysis for MDD2013 using FDRreg were also present in MDD2019 (original results without FDRreg) (**Table S8.4**). The statistical analyses yielded promising results: in most cases, the number of overlapping genes between the significant genes identified by the analysis for MDD2013 and the original significant results of MDD2019 was higher than that between the original significant results of MDD2013 and MDD2019 (9 out of 15 comparisons, **Table S8.4**). In the remaining comparisons, no overlapping genes were found before and after the analysis, making the comparison not applicable (**Table S8.4**).

In addition to internal validation, the drug cluster enrichment analysis for the genome-wide associated genes after FDRreg analysis of MDD2019 increased the prioritization of clusters specific to psychiatric disorders, compared to the genes from the original results (**Table 3** and **S8.9**). These included antipsychotics (FDR=8.93E-07), dopaminergic agents (FDR=1.52E-02), and hypnotics (FDR=2.57E-03). For the target MDD2013, the enrichment analysis showed an enriched signal for one of the three clusters in both before and after FDR regression analyses (hypnotics, FDR=2.43E-02 and 4.09E-02, respectively; **Table 3** and **S6.9**). Furthermore, the GO set enrichment analysis of the significant genes after the FDR analysis highlighted neuron/synapse related sets, including generation of neurons, regulation of nervous system development, synapse organization, and more (FDR<2.45E-10, **Table S8.5**). The significant genes from the genome-wide imputed gene expression study also highlighted neuron/synapse related pathways, such as neuronal system, synaptic adhesion-like molecules, and nicotine action pathway (FDR<4.61E-02, **Table S8.6**).

#### Bipolar disorder

For BPD, we selected two GWAS datasets with different sample size as our targets: BPD2012 and BPD2018 (**Table S1.1**). The genetic correlation analyses identified 21 co-variables significantly associated with BPD2012 and 27 associated with BPD2018 (**Table S9.1** and **Table S10.1**). Although the FDR regression analysis did not identify additional loci for the two BPD targets, it clearly discovered a significant higher number of associated genes for most of the gene-level analyses (FDR<0.01, **Table 1**).

The internal validation analysis suggested that the increased number of significant genes identified for BPD2012 through the FDR regression analysis are likely truly associated with the disorder. The number of overlapping genes between the significant genes from the FDR regression analysis of BPD2012 and the original significant results from BPD2018 was higher compared to that between the original significant results of BPD2012 and BPD2018 (14 out of 15 comparisons, **Table S10.4**). Additionally, the pathway and GO set enrichment analyses for the genome-wide significant genes after the FDR regression analysis showed significant enrichment signals in pathways and GO sets related to neurons and synapses (**Table S10.5**). Moreover, the individual drug and drug cluster enrichment analyses for BPD2018 identified a significantly higher number of enriched individual drugs (**Table S10.7**) and increased the prioritization of drug clusters specific to psychiatric disorders (**Table 3** and **S10.9**). These included antipsychotic, dopaminergic agents, anxiolytic, and hypnotics. For BPD2012, the enrichment analysis also significantly prioritized four types of drug clusters: anti-epileptics, antidepressants, antipsychotics, and dopaminergic agents (**Table 3** and **S9.9**).

#### Autism Spectrum Disorder

For ASD, we selected two GWAS datasets with different sample sizes as our targets: ASD2015 and ASD2019 (**Table S1.1**). The genetic correlation analyses revealed that 11 and 21 GWAS datasets were significantly associated with ASD2015 and ASD2019, respectively (p-value<0.05, **Table S11.1** and **Table S12.1**). The FDR regression analysis identified four additional genomic risk loci for ASD2019, increasing the total number of identified loci from 7 to 11 (**Table 1**). This method also discovered a significantly higher number of significant genes for all the gene-level association studies (FDR<0.01, **Table 1**). Furthermore, this analysis of ASD2015 identified two more significant genes for the imputed gene expression studies of the integrated results in 13 brain regions. Additionally, there were more significant genes identified in eight different brain regions (FDR<0.01, **Table 1**).

Due to the small number of significant signals in the original results from ASD2015 and ASD2019, we did not perform internal validation analysis. However, the pathway and GO set enrichment analyses for the genome-wide significant genes and significant associated imputed gene expression signals highlighted significant pathways and GO sets related to neurons and synapses. These included BDNF signaling pathway, motor neuron axon guidance, and GABAergic synapse (FDR<3.89E-02, **Table S12.4** and **S12.5**). Furthermore, compared to the original results, the drug cluster enrichment analysis for the genome-wide significant genes after the FDR regression analysis prioritized one psychiatry-related cluster, namely anti-epileptics, which ranked at the top 1 position (**Table S12.8**).

#### Attention Deficit/Hyperactivity Disorder and Suicide attempts

For ADHD, we selected two GWAS data-sets with different sample size as our targets: ADHD2016 and ADHD2019 (**Table S1.1**). In the genetic correlation analyses, we found that 10 co-variables were significantly associated with ADHD2016, while 28 co-variables associated with ADHD2019 (**Table S13.1** and **S14.1**). In Table 1, although the FDR regression analysis did not uncover additional genomic risk loci for ADHD2019, this method identified a significantly higher number of significant genes for all the gene-level association studies (FDR<0.01, **Table 1**). The pathway and GO set enrichment analyses for these genome-wide significant genes highlighted synapse or psychiatry related pathways and GO sets, such as twelve loci associated with ADHD (FDR=6.57E-17, **Table S14.4**), alcoholism (FDR=6.57E-15), and modification of postsynaptic structure (FDR=1.23E-02). Furthermore, compared to the original results, the drug cluster enrichment analysis for the genome-wide significant genes from ADHD2019 identified a significantly higher number of associated clusters (38 vs. 1; **Table S14.8**).

For SA, we selected three GWAS datasets as targets: SA in mental disorder (SA.iPSYCH), BPD (SA.BPD), and SCZ (SA.SCZ). These datasets were chosen because of varying underlying psychiatric disorders, or varying sample sizes (ranging from 4,629 to 50,624, **Table S1.1**). Due to the relatively low heritability of SA and moderate sample size, the genetic correlation analyses identified a limited number of associated co-variables for each SA target. Specifically, four associated co-variables were identified for SA.SCZ, nine for SA.BPD, and 18 for SA.iPSYCH (**Table S15.1, S16.1** and **S17.1**). Under these conditions, the FDR regression analysis only identified a slightly higher number of risk loci for the three SA targets (**Table 1**). The analysis of SA.iPSYCH identified one additional significant gene for the genome-wide associated study and more significant genes in five brain regions for imputed gene expression studies (amygdala, anterior cingulate cortex BA24, hypothalamus, nucleus accumbens basal ganglia, and substantia nigra, FDR<0.01, **Table 1**). Similarly, the analysis of SA.BPD determined four more significant genes for the imputed expression study of the integrated 13 brain regions, as well as more significant genes in the nucleus accumbens basal ganglia and spinal cord cervical regions (2 and 7, respectively, **Table 1**). For SA.SCZ, two additional significant genes were identified in the spinal cord cervical region for the imputed gene expression study. The pathway and GO sets enrichment analyses for these genome-wide significant genes highlighted many signals related to neurons and synapses. These include neurotransmitter release cycle, synaptic vesicle localization, and GABA synthesis, release, reuptake and degradation (FDR<1.40E-02, **Table S15.4**).

### Phenotypic study and experimental validation of ASD associated genes in embryonic brain development

To provide further support to the validity of the proposed FDR regression framework, we also examine the biological functions of additional associated genes for ASD. For the significantly associated genes for target ASD2019 dataset after FDR regression analysis, we identified thirty-one intersected ASD-associated genes across three different brain regions. Among them, a significant number of ten genes were found to exert biological functions based on the accumulating evidence in the OpenTarget database. To gain further insights into these genes, we first examined their expression level in different stages of central nerve system development and various organs in adults, using information from the mouse ENCODE transcriptome data in the NCBI database. We found that nine of them were expressed in both fetal and adult cortex, with exception of the Atg10 gene (**Fig. 2A**). Additionally, the Gabbr1 and Wdr73 genes obtained relatively high overall autism association scores (0.353 for Gabbr1 and 0.046 for Wdr73) from the OpenTarget database. Remarkably, the original results revealed no significant association between the *WDR73* gene and ASD2019 in any of the three brain regions (*P*>0.37). However, following the FDR regression analysis, the association reached statistical significance with a FDR of less than 0.05 (FDR<5.34E-03). As a result, we conducted IUE experiments to investigate the underlying mechanisms linking the two candidate genes to ASD during embryonic brain development. The main schematic diagram of the experimental setup was shown in **Fig. 2B**.

In the case of bicistronic constructs, we observed that two different shRNAs targeting each gene (shW1 and shW2 for WDR73; shG1 and shG2 for GABBR1) successfully knocked down the expression level of mouse Wdr73/Gabbr1 cDNA in 293T cells (**Fig. 2C-D**). Moving on to in vivo experiments, we examined the expression change of the two genes in NPCs through IUE at E16.5 rat brains. The positioning of GFP-positive (GFP+) cells showed that knockdown of Wdr73 resulted in a substantial reduction of cells in the ventricular zone (VZ)/subventricular zone (SVZ), accompanied by an increase in GFP+ cells in the cortical plate (CP) (**Fig. 2E and 2F**). This observation suggests that Wdr73 knockdown leads to a depletion of cells from the proliferative VZ/SVZ, indicating a potential role of Wdr73 in NPC proliferation and differentiation. To validate this hypothesis, we examined the overlap of the GFP+ electroporated cell population with different progenitor cell markers, including pax6 (marker for apical progenitor cells and radial glial cells) and Tbr2 (marker for intermediate or basal progenitors) (**Fig. 3A**). Indeed, Wdr73 knockdown resulted in a significant decrease in Pax6+ cell in the VZ/SVZ, indicating a reduced NPCs proliferation (**Fig. 3B**). Additionally, the increased proportion of cells double positive for pax6 and Tbr2 in GFP transfected cells suggested a premature differentiation of NPC cells (**Fig. 3C**). These findings indicated that Wdr73 may contribute to neuronal migration deficits, which is known to play an important role in the pathogenesis of ASD.

In brain sections with the Gabbr1 control vector, approximately 59% of GFP-positive cells migrated from the VZ/SVZ and intermediate zone (IZ) into the CP of the cortex (**Fig.2G and 2H**). However, in brain sections with Gabbr1 shRNAs, a significant number of GFP-positive neurons (∼59%) remained in the IZ and VZ/SVZ regions. These observations suggest that knockdown of the Gabbr1 leads to a suppression of radial migration.

## Discussion

The present study applied the FDR regression strategy on five types of psychiatric disorders and one psychiatric outcome, and identified additional genomic risk loci or associated genes/pathways for primary traits by integrating genetically correlated traits and biological annotations. Although various methods have been proposed to integrate results from univariate analyses, such as the Fisher’s combined p-value method and the more recent MTAG method [4–10, 12], they encounter inherent issues. The former method may lead to an inflated type I error when aggregating p-values of different but correlated phenotypes within the same cohort [4, 11]. On the other hand, the MTAG method is not efficient in generating the maximum FDR results when including more than 10 traits simultaneously, as observed in our pilot trial for estimating computational time. In contrast, the FDR regression analysis is able to handle GWAS datasets of over 100 traits (as covariables) within a few hours. This strategy only requires readily available GWAS summary statistics, such as the widely used UK Biobank [27–29]. Furthermore, the FDR regression model possesses inherent ability to control the FDR [11, 20]. Integrating numerous genetically correlated traits/phenotypes and biological annotations empowers the FDR regression model not only to identify additional associated risk loci/genes but also provide meaningful biological insights into these associated genes.

Compared to the original findings, the implementation of the FDR regression model strategy revealed a greater number of additional significant genes in most gene-level analyses (FDR<0.01, **Table 1**). Following the FDR regression analysis, we performed an internal validation analysis, which demonstrated higher-than-expected overlap in associated genes between small-scale GWAS analyses (SCZ2012, BPD2012 and MDD2013, **Table 2**) and their corresponding large-scale GWAS analyses (WAL.SCZ2018, BPD2018 and MDD2019) done subsequently. Furthermore, drug cluster enrichment analyses provided further validation by revealing stronger enrichment signals in psychiatry-specific drug clusters for the associated genes, thus underscoring their clinical significance compared to the original results with FDRreg (**Table 3**).

Notably, our study not only identified well-known genes and pathways associated with psychiatric disorders but also uncovered several novel genes and pathways in the large-scale studies (WAL.SCZ2018, BPD2018 and MDD2019). Taking WAL.SCZ2018 as an example, the pathway and GO sets enrichment analyses revealed that the genome-wide associated genes were significantly enriched in established neuron/synapse-related pathways and GO sets, such as neurotransmitter receptors, postsynaptic signal transmission, and generation of neurons (**Table S4**). Additionally, we observed significant enrichments in WNT signaling related pathways, including TCF dependent signaling in response to WNT and WNT signaling pathway and pluripotency (**Table S4.5**). While the WNT signaling has been extensively studied in cancer research, its role in regulating the function and structure of the adult nervous system has recently gained attention [30, 31]. Interestingly, dysregulated mRNA expression of Wnt-related genes in whole blood and plasma has been associated with attenuated canonical (beta-catenin-dependent) signaling in patients with SCZ and BPD [32]. Furthermore, the associated genes were found to be enriched in Rho GTPase activating proteins, including PKNs, formins, and KTN1. This family of activating proteins stimulates GTP hydrolysis, leading to the inactivation of the GTP-bound active form of Rho [33], which plays a crucial role in neuronal development processes [34–36]. Some members in this family, such as ARHGAP10 and ARHGAP18, have recently been associated with SCZ, [37, 38], while srGAP1, srGAP2, and srGAP3 have been linked to mental retardation, SCZ, and seizures [39]. Therefore, further investigation of these three activating proteins is warranted to explore their potential effects on psychiatry disorders.

In addition to identifying a significantly higher number of additional loci or genes associated with the primary traits of interest, we conducted IUE experiments to gain insights into the underlying mechanisms of these associations. Through a multiple-step pre-selection process, we focused on investigating the functional roles of the *Gabbr1* and *Wdr73* genes in the pathogenesis of ASD. Notably, the association between the *WDR73* gene in all three brain regions became statistically significant following the FDR regression analysis. This finding highlights the ability of our proposed method to effectively identify functionally important genes that might be overlooked in a conventional GWAS analysis. The IUE experiments conducted during embryonic brain development revealed that the *Wdr73* gene may play a crucial role in neuronal migration, while the *Gabbr1* gene is involved in radial migration. Collectively, our findings suggested that these two genes associated with ASD could impact distinct processes in embryonic brain development, which may ultimately contribute to behavioral changes after birth. Furthermore, these observations underscore the significance of early fetal brain development factors in the pathophysiology of ASD. It has been reported that the neurotransmitter receptor *GABBR1* is significantly downregulated in the brains of individuals with ASD compared to normal brains [40]. Additionally, one of expression quantitative trait loci (eQTLs) rs35828350, associated with ASD, has been found to downregulate *Wdr73* in the adult cortex [41]. Further exploration of the functional roles of other candidate genes involved in psychiatric traits or outcomes is warranted.

One limitation is the need to construct biological annotations if the FDR regression strategy is applied to other complex diseases. However, once the construction is completed, the matrix can be easily incorporated into the regression model, providing valuable information for the relevant primary diseases. In a similar vein, given the wide variety of biological annotations for SNPs/genes that are available, we have explored only a subset of such annotations and relevant databases, and exploration of other resources will be left as a future direction. We also note that the majority of GWAS studies are based on European populations, and the results may not be directly generalizable to other ethnicities. On the other hand, we have tried to include a few studies on East Asians. The FDRreg framework may also be useful to improve the statistical power of GWAS in non-European or minority ethnic groups, as shown in our application to the East Asian GWAS studies of SCZ. Due to the broad coverage of different psychiatric disorders and relatively large number of genes highlighted from this study, we only selected a few genes from the analysis of ASD for further experimental and mechanistic studies. Further experimental studies are required to elucidate the functional role of other genes uncovered in this study.

To conclude, the present study employed the FDR regression model and successfully identified a significantly higher number of additional genome risk loci or association genes for six common psychiatric disorders/outcomes. These additional association genes may have meaningful biological or clinical implications, as validated through internal validation analyses and drug cluster enrichment analyses. Furthermore, we performed further experimental work to reveal the functional importance of two additional genes in ASD. Compared to the original results, the additionally identified genes exhibited a stronger enriched signal in both well-known and novel pathways/GO sets. Importantly, this strategy holds substantial potential for the study of other complex human diseases, as it can leverage readily available GWAS summary statistics and may flexibly accommodate any useful biological annotations from other resources.

## Data Availability

All data produced in the present study are available upon reasonable request to the authors.

---

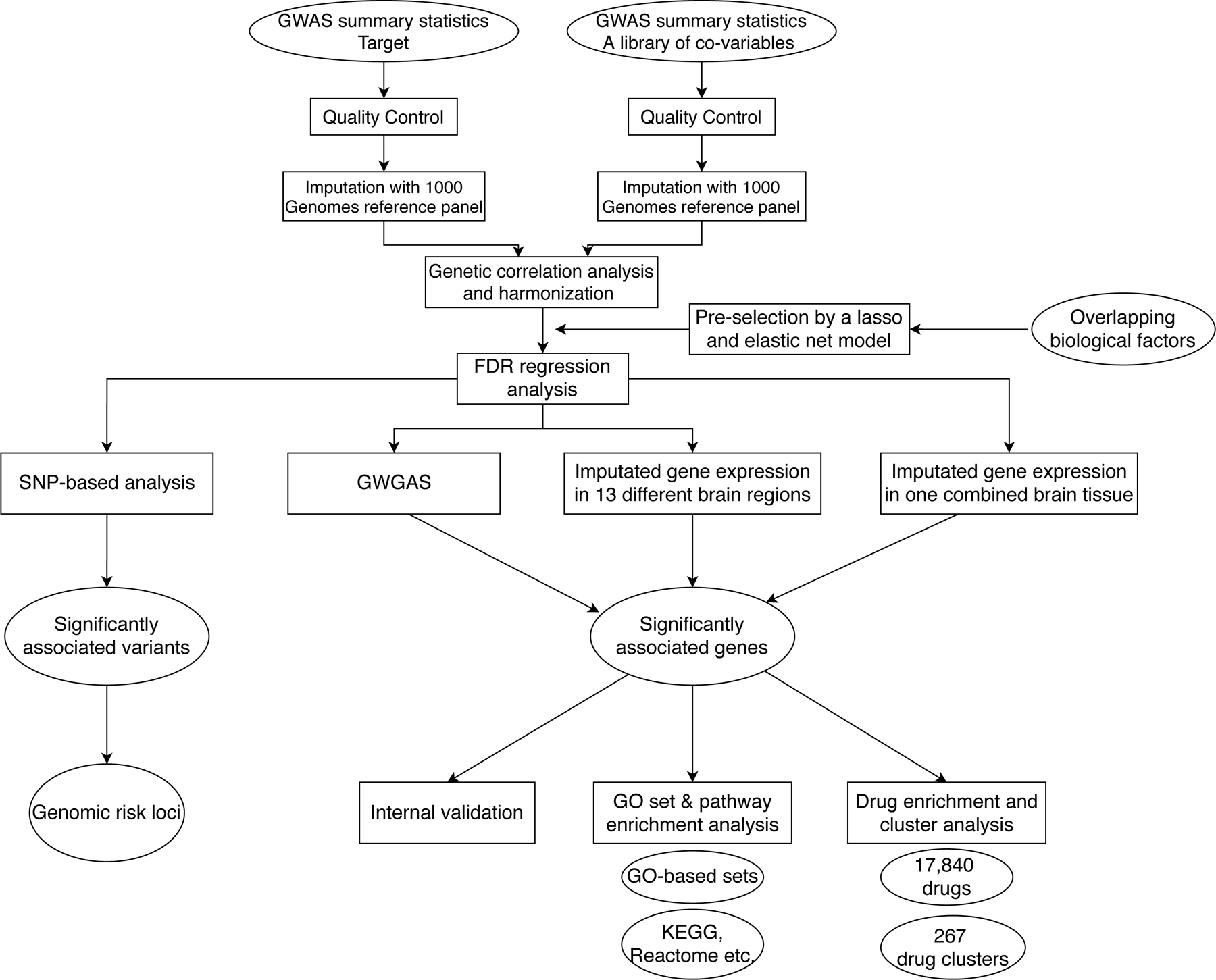

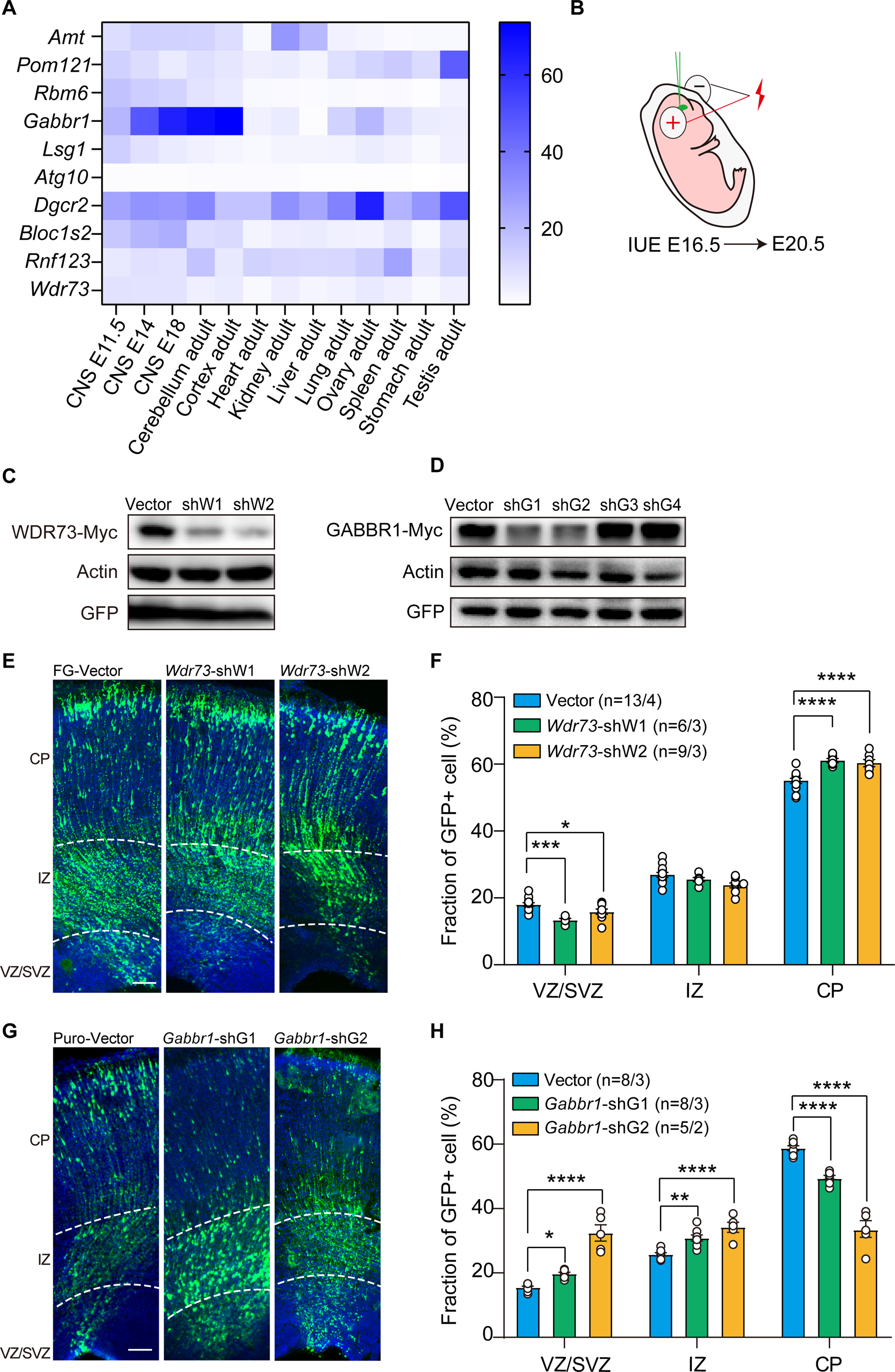

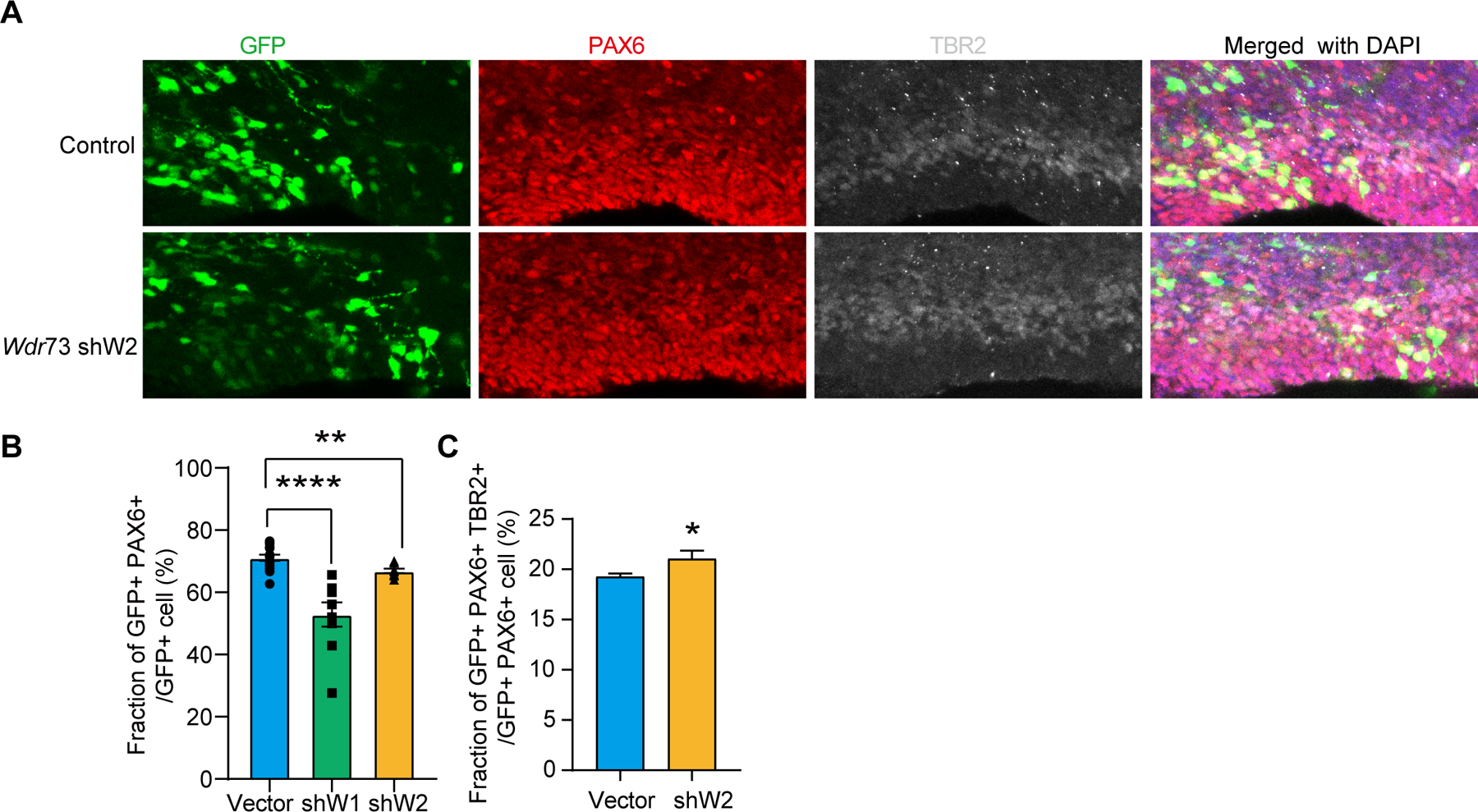

