## Supplementary Notes for "Discovering additional genetic loci associated with six psychiatric disorders/traits *via* FDR regression model leveraging external genetic and biological data"

**Supplementary Materials**

1. **Functional annotations for mapping genes in three databases**

For annotations of mapping gene, we selected six aspects of biological information from the database for annotation, visualization and integrate discovery (DAVID, v6.8), that is, expression-specific tissues, associated diseases, relevant pathways, transcription factor binding site (TFBS), biological processes and biological interactions (Huang da et al., 2009). For expression-specific tissues, a score of ‘1’ would assign to one gene if any of following keywords present: brain, cortex, cerebellar, cerebellum, putamen, caudate, nucleus accumbens, hippocampus, amygdala, spinal cord, hypothalamus, substantia nigra, synapse and synaptic. The other genes would be scored of ‘0’. As for associated diseases, a score of ‘2’ would assign to one gene if any of follow-up keywords present: schizo, depress, bipolar, autis, attention deficit, alcohol, circadian, drug abuse. A score of ‘1’ would assign to one gene that associated with any other disease and a score of ‘0’ would assign to the remaining genes. Regarding relevant pathways, one gene would be assigned score of ‘2’ if it was involved in brain relevant pathways, including keywords like neuro, alcohol, synapse, synaptic, serotonergic and gabaergic. A score of ‘1’ would assign to one gene if it was involved in any biological pathway and the others would be assigned score of ‘0’. With regard to TFBS, a score of ‘1’ would assign to one gene if it has more than one TFBS and the others would be assigned score of ‘0’. For biological processes, a score of ‘2’ would be assign to one gene if any of the following keywords present: brain, dopamine, serotonin, neuro, tryptophan, epinephrine, histamine, norepinephrine, circadian, cortex, cerebeller, cerebellum, putamen, caudate, nucleus accumbens, hippocampus, amygdala, spinal cord, hypothalamus, substantia nigra, schizo, depress, bipolar, autis, attention deficit, alcohol, synapse and synaptic. A score of ‘1’ would assign to one gene if it was involved in any other biological process. The remaining genes would be scored of ‘0’. As for biological interaction, a score of ‘2’ would assign to one gene if any of following keywords present: brain, dopamine, serotonin, neuro, tryptophan, epinephrine, histamine, norepinephrine, circadian, synapse and synaptic. A score of ‘1’ would assign to one gene if it had any other biological interaction and the others would be assigned score of ‘0’.

Regarding the Open Target database, the original score calculated by its algorithm would be used in the present study (Carvalho-Silva et al., 2018). For the denovo database, a score of ‘1’ would be assigned to one gene if it had any denovo mutation that appeared in either SCZ or ASD patients. The others would be scored of ‘0’ (denovo-db, Seattle, WA (URL: denovo-db.gs.washington.edu) [02-2020 accessed]).

**2. Phenotypic study of candidate genes in embryonic brain development**

Antibodies used for immunohistochemistry were as follows: rabbit anti-PAX6 (abcam), chicken anti-TBR1 (Millipore). The following antibodies were used for western blot: mouse anti-Myc (CST), rabbit anti-Actin (Servicebio), rabbit anti-GFP (abcam).

Mouse WDR73-Myc or GABBR1-Myc was created by cloning WDR73 or GABBR1 cDNA sequence into the pcDNA3.1 myc-His A vector. Small hairpin constructs were cloned into either FUGW-H1-Scrambled vector or pLKO.1-TRC-copGFP-2A-PURO vector. Wdr73 shRNA sequence was cloned into FUGW-H1-Scrambled vector. WDR73 small hairpin constructs containing the following target sequences were used: shW1: 5’- GGGACTTCAAAGTACGCCA-3’;

shW2: 5’- GGCTATTTCAGGCTTTGAT-3’.

Gabbr1 shRNA sequence was cloned into pLKO.1-TRC-copGFP-2A-PURO vector. GABBR1 small hairpin constructs containing the following target sequences were used:

shG1: 5’- CCCGGATGTGGAACCTTATTG-3’;

shG2: 5’- TCGGAAGGTTGCCAGATTATA-3’;

shG3: 5’- GCACCGAACCATTGAGACTTT-3’;

shG4: 5’- TGAACCCTGCTTGGAGCTATT-3’.

**3. Brief descriptions of 17 supplementary tables.**

**Table S1.** Targets and co-variables for FDR regression analysis and internal validation.

**S1.1.** Included 16 GWAS summary statistics as targets for FDR regression analysis.

**S1.2.** Included 42 GWAS summary statistics as co-variables for FDR regression analysis.

**S1.3.** Internal validation of detection capacity and accuracy for FDR regression model.

**Table S2.** Overall results for SCZ2012 in psychiatric genomics consortium.

**S2.1.** Genetic correlation analyses between target phenotype and a library of 42 pre-defined related phenotypes.

**S2.2.** Contribution of those associated features in FDR regression analyses (SNP-based, gene-association-based and imputed-gene-expression-based).

**S2.3.** Contribution of those related features in FDR regression analysis models in 13 individual brain regions (imputed-gene-expression-based).

**S2.4.** Identification of genomic risk loci for significant SNPs before or after FDR regression analysis.

**S2.5.** Significant genes identified by genome-wide gene-based association study (GWGAS) in MAGMA program.

**S2.6.** Significant genes identified by genome-wide imputed gene expression association study in SMultiXcan program. pvalue: nominal p-value that generated by S-MultiXcan; P(adj): caculated by R function p.adjust for nominal p-value; bio.FDR.the: caculated by R funtion FDRreg with theoretical null type.

**S2.7.** Individual drug enrichment analysis for significant genes with or without FDR regression analysis (gene-association-based).

**S2.8.** Individual drug enrichment analysis for significant genes with or without FDR regression analysis (imputed-gene-expression-based).

**S2.9.** Drug cluster enrichment analysis for significant genes with or without FDR regression analysis (gene-association-based).

**S2.10.** Drug cluster enrichment analysis for significant genes with or without FDR regression analysis (imputed-gene-expression-based).

**Table S3.** Overall results for SCZ2014 in psychiatric genomics consortium.

**S3.1.** Genetic correlation analyses between target phenotype and a library of 42 pre-defined related phenotypes.

**S3.2.** Contribution of those associated features in FDR regression analyses (SNP-based, gene-association-based and imputed-gene-expression-based).

**S3.3.** Contribution of those related features in FDR regression analysis models in 13 individual brain regions (imputed-gene-expression-based).

**S3.4.** Internal validation based on significant genes from small-size and big-size GWAS.

**S3.5.** Identification of genomic risk loci for significant SNPs before or after FDR regression analysis.

**S3.6.** Significant genes identified by genome-wide gene-based association study (GWGAS) in MAGMA program.

**S3.7.** Significant genes identified by genome-wide imputed gene expression association study in SMultiXcan program.

**S3.8.** Individual drug enrichment analysis for significant genes with or without FDR regression analysis (gene-association-based).

**S3.9.** Individual drug enrichment analysis for significant genes with or without FDR regression analysis (imputed-gene-expression-based).

**S3.10.** Drug cluster enrichment analysis for significant genes with or without FDR regression analysis (gene-association-based)

**S3.11.** Drug cluster enrichment analysis for significant genes with or without FDR regression analysis (imputed-gene-expression-based).

Genetic correlation analyses between target phenotype and a library of 42 pre-defined related phenotypes.

**Table S4.** Overall results for SCZ2018 from Walter’s group.

**S4.1.** Genetic correlation analyses between target phenotype and a library of 42 pre-defined related phenotypes.

**S4.2.** Contribution of those associated features in FDR regression analyses (SNP-based, gene-association-based and imputed-gene-expression-based).

**S4.3.** Contribution of those related features in FDR regression analysis models in 13 individual brain regions (imputed-gene-expression-based).

**S4.4.** Internal validation based on significant genes from small-size and big-size GWAS.

**S4.5.** Pathway&GO set enrichment analysis for significant genes from whole-genome gene-based analysis using the MAGMA program.

**S4.6.** Pathway&GO set enrichment analysis for significant genes from whole-genome imputed gene expression association analysis using the Smultixcan program.

**S4.7.** Individual drug enrichment analysis for significant genes with or without FDR regression analysis (gene-association-based).

**S4.8.** Individual drug enrichment analysis for significant genes with or without FDR regression analysis (imputed-gene-expression-based).

**S4.9.** Drug cluster enrichment analysis for significant genes with or without FDR regression analysis (gene-association-based).

**S4.10.** Drug cluster enrichment analysis for significant genes with or without FDR regression analysis (imputed-gene-expression-based).

**S4.11.** Identification of genomic risk loci for significant SNPs before or after FDR regression analysis.

**S4.12.** Significant genes identified by genome-wide gene-based association study (GWGAS) in MAGMA program.

**S4.13.** Significant genes identified by genome-wide imputed gene expression association study in SMultiXcan program.

**Table S5.** Overall results for SCZ2019 in eastern Asian population.

**S5.1.** Genetic correlation analyses between target phenotype and a library of 42 pre-defined related phenotypes.

**S5.2.** Contribution of those associated features in FDR regression analyses (SNP-based, gene-association-based and imputed-gene-expression-based).

**S5.3.** Contribution of those related features in FDR regression analysis models in 13 individual brain regions (imputed-gene-expression-based).

**S5.4.** Identification of genomic risk loci for significant SNPs before or after FDR regression analysis.

**S5.5.** Significant genes identified by genome-wide gene-based association study (GWGAS) in MAGMA program.

**S5.6.** Significant genes identified by genome-wide imputed gene expression association study in SMultiXcan program.

**S5.7.** Individual drug enrichment analysis for significant genes with or without FDR regression analysis (gene-association-based).

**S5.8.** Individual drug enrichment analysis for significant genes with or without FDR regression analysis (imputed-gene-expression-based).

**S5.9.** Drug cluster enrichment analysis for significant genes with or without FDR regression analysis (gene-association-based).

**S5.10.** Drug cluster enrichment analysis for significant genes with or without FDR regression analysis (imputed-gene-expression-based).

**Table S6.** Overall results for MDD2013 in psychiatric genomics consortium.

**S6.1.** Genetic correlation analyses between target phenotype and a library of 42 pre-defined related phenotypes.

**S6.2.** Contribution of those associated features in FDR regression analyses (SNP-based, gene-association-based and imputed-gene-expression-based).

**S6.3.** Contribution of those related features in FDR regression analysis models in 13 individual brain regions (imputed-gene-expression-based).

**S6.4.** Identification of genomic risk loci for significant SNPs before or after FDR regression analysis.

**S6.5.** Significant genes identified by genome-wide gene-based association study (GWGAS) in MAGMA program.

**S6.6.** Significant genes identified by genome-wide imputed gene expression association study in SMultiXcan program.

**S6.7.** Individual drug enrichment analysis for significant genes with or without FDR regression analysis (gene-association-based).

**S6.8.** Individual drug enrichment analysis for significant genes with or without FDR regression analysis (imputed-gene-expression-based).

**S6.9.** Drug cluster enrichment analysis for significant genes with or without FDR regression analysis (gene-association-based).

**S6.10.** Drug cluster enrichment analysis for significant genes with or without FDR regression analysis (imputed-gene-expression-based).

**Table S7.** Overall results for the CONVERGE study of MDD.

**S7.1.** Genetic correlation analyses between target phenotype and a library of 42 pre-defined related phenotypes.

**S7.2.** Contribution of those associated features in FDR regression analyses (SNP-based, gene-association-based and imputed-gene-expression-based).

**S7.3.** Contribution of those related features in FDR regression analysis models in 13 individual brain regions (imputed-gene-expression-based).

**S7.4.** Identification of genomic risk loci for significant SNPs before or after FDR regression analysis.

**S7.5.** Significant genes identified by genome-wide gene-based association study (GWGAS) in MAGMA program.

**S7.6.** Significant genes identified by genome-wide imputed gene expression association study in SMultiXcan program.

**S7.7.** Individual drug enrichment analysis for significant genes with or without FDR regression analysis (gene-association-based).

**S7.8.** Individual drug enrichment analysis for significant genes with or without FDR regression analysis (imputed-gene-expression-based).

**S7.9.** Drug cluster enrichment analysis for significant genes with or without FDR regression analysis (gene-association-based).

**S7.10.** Drug cluster enrichment analysis for significant genes with or without FDR regression analysis (imputed-gene-expression-based).

**Table S8.** Overall results for MDD2019 in psychiatric genomics consortium.

**S8.1.** Genetic correlation analyses between target phenotype and a library of 42 pre-defined related phenotypes.

**S8.2.** Contribution of those associated features in FDR regression analyses (SNP-based, gene-association-based and imputed-gene-expression-based).

**S8.3.** Contribution of those related features in FDR regression analysis models in 13 individual brain regions (imputed-gene-expression-based).

**S8.4.** Internal validation based on significant genes from small-size and big-size GWAS.

**S8.5.** Pathway&GO set enrichment analysis for significant genes from whole-genome gene-based analysis using the MAGMA program.

**S8.6.** Pathway&GO set enrichment analysis for significant genes from whole-genome imputed gene expression association analysis using the Smultixcan program.

**S8.7.** Individual drug enrichment analysis for significant genes with or without FDR regression analysis (gene-association-based).

**S8.8.** Individual drug enrichment analysis for significant genes with or without FDR regression analysis (imputed-gene-expression-based).

**S8.9.** Drug cluster enrichment analysis for significant genes with or without FDR regression analysis (gene-association-based).

**S8.10.** Drug cluster enrichment analysis for significant genes with or without FDR regression analysis (imputed-gene-expression-based).

**S8.11.** Identification of genomic risk loci for significant SNPs before or after FDR regression analysis.

**S8.12.** Significant genes identified by genome-wide gene-based association study (GWGAS) in MAGMA program.

**S8.13.** Significant genes identified by genome-wide imputed gene expression association study in SMultiXcan program.

**Table S9.** Overall results for BD2012 in psychiatric genomics consortium.

**S9.1.** Genetic correlation analyses between target phenotype and a library of 42 pre-defined related phenotypes.

**S9.2.** Contribution of those associated features in FDR regression analyses (SNP-based, gene-association-based and imputed-gene-expression-based).

**S9.3.** Contribution of those related features in FDR regression analysis models in 13 individual brain regions (imputed-gene-expression-based).

**S9.4.** Identification of genomic risk loci for significant SNPs before or after FDR regression analysis.

**S9.5.** Significant genes identified by genome-wide gene-based association study (GWGAS) in MAGMA program.

**S9.6.** Significant genes identified by genome-wide imputed gene expression association study in SMultiXcan program.

**S9.7.** Individual drug enrichment analysis for significant genes with or without FDR regression analysis (gene-association-based).

**S9.8.** Individual drug enrichment analysis for significant genes with or without FDR regression analysis (imputed-gene-expression-based).

**S9.9.** Drug cluster enrichment analysis for significant genes with or without FDR regression analysis (gene-association-based).

**S9.10.** Drug cluster enrichment analysis for significant genes with or without FDR regression analysis (imputed-gene-expression-based).

**Table S10.** Overall results for BD2018 in psychiatric genomics consortium.

**S10.1.** Genetic correlation analyses between target phenotype and a library of 42 pre-defined related phenotypes.

**S10.2.** Contribution of those associated features in FDR regression analyses (SNP-based, gene-association-based and imputed-gene-expression-based).

**S10.3.** Contribution of those related features in FDR regression analysis models in 13 individual brain regions (imputed-gene-expression-based).

**S10.4.** Internal validation based on significant genes from small-size and big-size GWAS.

**S10.5.** Pathway&GO set enrichment analysis for significant genes from whole-genome gene-based analysis using the MAGMA program.

**S10.6.** Pathway&GO set enrichment analysis for significant genes from whole-genome imputed gene expression association analysis using the Smultixcan program.

**S10.7.** Individual drug enrichment analysis for significant genes with or without FDR regression analysis (gene-association-based).

**S10.8.** Individual drug enrichment analysis for significant genes with or without FDR regression analysis (imputed-gene-expression-based).

**S10.9.** Drug cluster enrichment analysis for significant genes with or without FDR regression analysis (gene-association-based).

**S10.10.** Drug cluster enrichment analysis for significant genes with or without FDR regression analysis (imputed-gene-expression-based).

**S10.11.** Identification of genomic risk loci for significant SNPs before or after FDR regression analysis.

**S10.12.** Significant genes identified by genome-wide gene-based association study (GWGAS) in MAGMA program.

**S10.13.** Significant genes identified by genome-wide imputed gene expression association study in SMultiXcan program.

**Table S11.** Overall results for ASD2015 in psychiatric genomics consortium.

**S11.1.** Genetic correlation analyses between target phenotype and a library of 37 pre-defined related phenotypes.

**S11.2.** Contribution of those associated features in FDR regression analyses (SNP-based, gene-association-based and imputed-gene-expression-based).

**S11.3.** Contribution of those related features in FDR regression analysis models in 13 individual brain regions (imputed-gene-expression-based).

**S11.4.** Significant genes identified by genome-wide gene-based association study (GWGAS) in MAGMA program.

**S11.5.** Significant genes identified by genome-wide imputed gene expression association study in SMultiXcan program.

**Table S12.** Overall results for ASD2019 in the integrative psychiatric research project.

**S12.1.** Genetic correlation analyses between target phenotype and a library of 37 pre-defined related phenotypes.

**S12.2.** Contribution of those associated features in FDR regression analyses (SNP-based, gene-association-based and imputed-gene-expression-based).

**S12.3.** Contribution of those related features in FDR regression analysis models in 13 individual brain regions (imputed-gene-expression-based).

**S12.4.** Pathway&GO set enrichment analysis for significant genes from whole-genome gene-based analysis using the MAGMA program.

**S12.5.** Pathway&GO set enrichment analysis for significant genes from whole-genome imputed gene expression association analysis using the Smultixcan program.

**S12.6.** Individual drug enrichment analysis for significant genes with or without FDR regression analysis (gene-association-based).

**S12.7.** Individual drug enrichment analysis for significant genes with or without FDR regression analysis (imputed-gene-expression-based).

**S12.8.** Drug cluster enrichment analysis for significant genes with or without FDR regression analysis (gene-association-based).

**S12.9.** Drug cluster enrichment analysis for significant genes with or without FDR regression analysis (imputed-gene-expression-based).

**S12.10.** Identification of genomic risk loci for significant SNPs before or after FDR regression analysis.

**S12.11.** Significant genes identified by genome-wide gene-based association study (GWGAS) in MAGMA program.

**S12.12.** Significant genes identified by genome-wide imputed gene expression association study in SMultiXcan program.

**Table S13.** Overall results for ADHD2016 in the EAGLE study.

**S13.1.** Genetic correlation analyses between target phenotype and a library of 37 pre-defined related phenotypes.

**S13.2.** Contribution of those associated features in FDR regression analyses (SNP-based, gene-association-based and imputed-gene-expression-based).

**S13.3.** Contribution of those related features in FDR regression analysis models in 13 individual brain regions (imputed-gene-expression-based).

**S13.4.** Significant genes identified by genome-wide gene-based association study (GWGAS) in MAGMA program.

**S13.5.** Significant genes identified by genome-wide imputed gene expression association study in SMultiXcan program.

**Table S14.** Overall results for ADHD2019 in the integrative psychiatric research project.

**S14.1.** Genetic correlation analyses between target phenotype and a library of 42 pre-defined related phenotypes.

S14.2. Contribution of those associated features in FDR regression analyses (SNP-based, gene-association-based and imputed-gene-expression-based).

S14.3. Contribution of those related features in FDR regression analysis models in 13 individual brain regions (imputed-gene-expression-based).

S14.4. Pathway&GO set enrichment analysis for significant genes from whole-genome gene-based analysis using the MAGMA program.

S14.5. Pathway&GO set enrichment analysis for significant genes from whole-genome imputed gene expression association analysis using the Smultixcan program.

S14.6. Individual drug enrichment analysis for significant genes with or without FDR regression analysis (gene-association-based).

S14.7. Individual drug enrichment analysis for significant genes with or without FDR regression analysis (imputed-gene-expression-based).

S14.8. Drug cluster enrichment analysis for significant genes with or without FDR regression analysis (gene-association-based)

S14.9. Drug cluster enrichment analysis for significant genes with or without FDR regression analysis (imputed-gene-expression-based).

S14.10. Identification of genomic risk loci for significant SNPs before or after FDR regression analysis.

S14.11. Significant genes identified by genome-wide gene-based association study (GWGAS) in MAGMA program.

S14.12. Significant genes identified by genome-wide imputed gene expression association study in SMultiXcan program.

**Table S15.** Overall results for SA in mental disorders in the integrative psychiatric research project.

**S15.1.** Genetic correlation analyses between target phenotype and a library of 37 pre-defined related phenotypes.

**S15.2.** Contribution of those associated features in FDR regression analyses (SNP-based, gene-association-based and imputed-gene-expression-based).

**S15.3.** Contribution of those related features in FDR regression analysis models in 13 individual brain regions (imputed-gene-expression-based).

**S15.4.** Pathway&GO set enrichment analysis for significant genes from whole-genome gene-based analysis using the MAGMA program.

**S15.5.** Significant genes identified by genome-wide gene-based association study (GWGAS) in MAGMA program.

**S15.6.** Significant genes identified by genome-wide imputed gene expression association study in SMultiXcan program.

**Table S16.** Overall results for SA in bipolar disorder in psychiatric genomics consortium.

**S16.1.** Genetic correlation analyses between target phenotype and a library of 37 pre-defined related phenotypes.

**S16.2.** Contribution of those associated features in FDR regression analyses (SNP-based, gene-association-based and imputed-gene-expression-based).

**S16.3.** Contribution of those related features in FDR regression analysis models in 13 individual brain regions (imputed-gene-expression-based).

**S16.4.** Pathway&GO set enrichment analysis for significant genes from whole-genome imputed gene expression association analysis using the Smultixcan program.

**S16.5.** Significant genes identified by genome-wide gene-based association study (GWGAS) in MAGMA program.

**S16.6.** Significant genes identified by genome-wide imputed gene expression association study in SMultiXcan program.

**Table S17.** Overall results for SA in schizophrenia in psychiatric genomics consortium.

**S17.1.** Genetic correlation analyses between target phenotype and a library of 37 pre-defined related phenotypes.

**S17.2.** Contribution of those associated features in FDR regression analyses (SNP-based, gene-association-based and imputed-gene-expression-based).

**S17.3.** Contribution of those related features in FDR regression analysis models in 13 individual brain regions (imputed-gene-expression-based).

**S17.4.** Significant genes identified by genome-wide gene-based association study (GWGAS) in MAGMA program.

**S17.5.** Significant genes identified by genome-wide imputed gene expression association study in SMultiXcan program.
